# Detection of SARS-CoV-2 neutralizing antibodies with a cell-free PCR assay

**DOI:** 10.1101/2020.05.28.20105692

**Authors:** Kenneth Danh, Donna Grace Karp, Peter V. Robinson, David Seftel, Mars Stone, Graham Simmons, Anil Bagri, Martin Schreiber, Andreas Buser, Andreas Holbro, Manuel Battegay, Laurence M. Corash, Carl Hanson, Cheng-ting Tsai

## Abstract

Severe acute respiratory syndrome coronavirus 2 (SARS-CoV-2) has led to more than 4 million confirmed infections worldwide and over 300,000 deaths. While Remdesivir has recently received FDA emergency use authorization for treatment of SARS-CoV-2 infection, convalescent plasma (CP) with high titers of SARS-CoV-2 neutralizing antibodies (NAbs) from recovered donors remains a promising and widely accessible method to mitigate severe disease symptoms. Here, we describe the development and validation of a cell-free neutralization PCR assay using SARS-CoV-2 spike protein S1 and human ACE2 receptor-DNA conjugates. By comparing with samples collected prior to the outbreak, we confirmed that NAbs were specifically detected in COVID-19 cases. Using our unique assay, the NAb signals are detectable as early as 10 days after onset of symptoms and continue to rise, plateauing after 18 days. Notably, we showed that the use of a licensed pathogen reduction technology to inactivate potentially contaminating infectious pathogens in CP did not alter NAb signals, paving a path to safely administer effective CP therapies. The described neutralization PCR assay can serve as a qualification tool to easily identify suitable CP donors of a potentially lifesaving therapy. In addition, this assay tool is readily deployable in standard laboratories with biosafety level 2 capability, and can yield results within 2-3 hrs. This advancement can facilitate research on factors driving diverse COVID-19 disease manifestations, help evaluate the impact of various CP processing protocols on CP therapeutic efficacy and assist in accelerating vaccine efficacy assessment.

## INTRODUCTION

The current epidemic of COVID-19 (novel coronavirus disease-2019) caused by SARS-CoV-2 has propagated globally at an unprecedented speed. It has led to more than 4 million confirmed infections worldwide and over 300,000 deaths. COVID-19 disease is particularly challenging in that there are few broadly effective and specialized treatments to contain the disease and mitigate severe symptoms^1,2^. Convalescent plasma (CP) has garnered strong interest since it is readily available from recovered patients and has been used with some efficacy in past pandemics, including the 2009-2010 H1N1 influenza and the 2013 West Africa Ebola outbreak^3^.

The primary mechanism of action of CP is through infusing neutralizing antibodies (NAbs) harvested from recovered patients to disrupt viral entry into host cells in acutely infected recipient patients^3-5^. The identification of suitable donors for prompt administration of CP remains a major unmet need for the effective clinical deployment of CP. Current serological assays simply detect the interaction of antibody with cognate viral antigens. Reliance on this interaction, while sufficient for diagnosis, is not indicative of neutralization capacity, and may lead to therapeutically ineffective CP without active NAb components. However, current assays that competently assess NAbs are time-consuming and labor intensive, causing a significant bottleneck to widespread administration of high-quality CP^6,7^.

The virus plaque reduction neutralization test (PRNT) is the current gold standard assay for NAbs^6^. However, PRNT’s reliance on large quantities of infectious SARS-CoV-2 virions limits the use of this potentially hazardous and time-consuming assay to relatively few well-resourced institutes with biosafety level 3 (BSL3) laboratories. Modifications have been implemented to improve the safety profile of the PRNT, but its fundamental reliance on cell culturing requires dedicated clean room facilities and several days of observation for measuring impact on cell death. For instance, pseudovirus neutralization assays port sections of the virus in question into benign viral hosts to allow for a safer approximation of PRNT, but are still reliant on slow and expensive cell-based methods^6^. Therefore, the creation of a high-throughput, rapid and easily-implementable assay for NAbs for CP therapy remains a high priority.

In this study, we constructed and validated a cell-free assay to measure NAbs using COVID-19 and control patient samples. This assay was inspired in part by our previous work with the antibody detection by agglutination PCR (ADAP) methodology that has been successfully used to develop and validate ultrasensitive and highly specific assays for wide variety of infections and autoimmune diseases, including HIV, food allergy and type 1 diabetes^8-10^. Notably, we used this cell-free assay to characterize antibody activity in samples from CP used for patient transfusions.

## METHODS

### Materials

The SARS-CoV-2 spike protein (S1) containing amino acids 1-674 with an Fc-tag at the C-terminus (#31806) expressed in HEK293 cells was purchased from the Native Antigen Company (Oxford, United Kingdom). The SARS-CoV-2 spike protein receptor binding domain (RBD) containing amino acids 319-541 with an Fc-tag at the C-terminus (#40592-V02H) and the human receptor angiotensin-converting enzyme 2 (ACE2) protein containing amino acids 1-740 with an Fc-tag at the C-terminus (10108- H05H) expressed in HEK293 cells were obtained from Sino Biologicals (Beijing, China). Oligonucleotides used in the study were custom ordered from Integrated DNA Technologies (Coralville, IA). Platinum Taq polymerase (#10966026), SYBR qPCR 2X master mix (#4385610), Dithiothreitol (DTT #202090) and sulfo-SMCC (#22122) were purchased from Thermo Fisher (Waltham, MA). DNA ligase (#A8101) was purchased from LGC (Teddington, United Kingdom). Other reagents are detailed in the method sections as appropriate.

### Human specimens used in the study

Blood specimens from SARS-CoV-2 RNA positive individuals were obtained from various sources. Two serum samples were obtained from COVID-19 patients from the Oregon Health Sciences University Hospital (OHSU), Portland, OR. These were sourced from discarded clinical laboratory specimens exempted from informed consent and IRB approval under condition of patient anonymity. Three plasma samples were obtained from COVID-19 convalescent outpatient plasma donors of the University Hospital Basel, University of Basel, Basel, Switzerland with informed consent. These donors were screened in compliance with Swiss regulations on blood donation and approved as plasma donors in accordance with national regulations. Three convalescent COVID-19 patients were recruited to donate serum samples for the study with informed consent under Enable Biosciences IRB #20180015 (approved by Western IRB). Twenty-five remnant serum samples sourced from discarded clinical laboratory specimens were obtained from the California Department of Public Health. Forty-seven serum samples collected prior to the outbreak from blood donors and individuals with other viral or bacterial infection were purchased from commercial biobanks. All specimens collected outside of Enable Biosciences clinical network were received as de-identified specimens.

### Synthesis of protein-DNA conjugates for neutralization assay

For S1, RBD and ACE2-DNA conjugates, the proteins were buffer exchanged in reaction buffers (55 mM sodium phosphate, 150 mM sodium chloride, 20 mM EDTA, pH 7.2) to make 1 mg/mL solutions. A 1 μL solution of 8mM sulfo-SMCC was added to 10 μL of each protein solution. The reaction mixtures were incubated at room temperature for 2 hours. Thiolated DNA was suspended in reaction buffers to 100 μM. A 3 μL solution of thiolated DNA solution and 4 μL of 100 mM solution of DTT were mixed to reduce dimerized thiolated DNA to monomer forms. The solution was then incubated at 37 °C for 1 hour. The excess sulfo-SMCC in protein mixtures and DTT in thiolated-DNA were removed by 7K MWCO Zeba spin column (Thermo Fischer, Waltham, MA). The thiolated DNA and protein solutions were then pooled and incubated overnight at 4 °C. Finally, protein-DNA conjugates were purified by 30 kDa MWCO filter (Millipore, Burlington, MA). Conjugate concentrations were determined by BCA assay (Thermo Fischer). Conjugation efficiencies were analyzed by SDS-PAGE and silver staining as described previously^8-10^. DNA-to-protein ratios of the conjugates were estimated by UV-VIS absorption and typically fell in the range of 2- to-1. Protein-DNA conjugates were stored at 4 °C for short-term usage or aliquoted for long-term storage at −80 °C.

### Neutralization PCR assay

The cell-free neutralization assay detects NAbs by observing the disruption of interaction between the S1 protein and the ACE2 receptor (**Fig. 1**). Briefly, 1 μL of blood sample (e.g. serum, plasma) was incubated with 2 μL of 1 femtomole of S1-DNA conjugate mixtures at 37 °C for 30 min. Then, 2μL of 1 femtomole of ACE2-DNA conjugate mixtures was added and incubated at 37 °C for another 30 min. The neutralizing antibodies in the specimen will engage with S1-DNA conjugate in step 1 to decrease S1-DNA binding with ACE2-DNA in step 2. To quantify the degree of competition, 115 μl of ligation mix (20 mM Tris, 50 mM KCl, 20 mM MgCl2, 20 mM DTT, 25 μM NAD, 0.025 U/μl ligase, 100 nM connector) was added and incubated at 30 °C for 15 min. Then, 25 μL of ligated solution was mixed with the PCR master mix that contained primer pairs and polymerase for amplification under standard thermocycling conditions (95 °C for 10 min, 95 °C for 15 sec, 56 °C for 30 sec, 13 cycles). The pre-amplified products were quantified in a 96-well qPCR plate. SYBR green-based qPCR was performed on a Bio-Rad CFX96 real-time PCR detection system (95 °C for 10 min, 95 °C for 30 sec, 56 °C for 1 min, 40 cycles). As a result of competition, specimens harboring high quantities of NAbs will have less amplifiable DNA, thus a weaker qPCR signal (higher Ct). In contrast, samples without neutralizing antibodies will benefit from the strong binding between S1 and ACE2 proteins to generate a large amount of DNA amplicons, thus a stronger qPCR signal (lower Ct). Instead of using the common cycle threshold (Ct) as a readout, the neutralization PCR assay readout ΔCt is defined as the Ct value of the actual sample minus the Ct of a blank control. The magnitude of the ΔCt is proportional to the loss of amplicon concentration in the PCR plate well, which in turn is proportional to the amount of neutralizing antibody present in the sample. The ΔCt offers significant reproducibility since the subtraction of the blank control Ct and the sample Ct cancels out any potential drift across runs.

**Figure 1.**
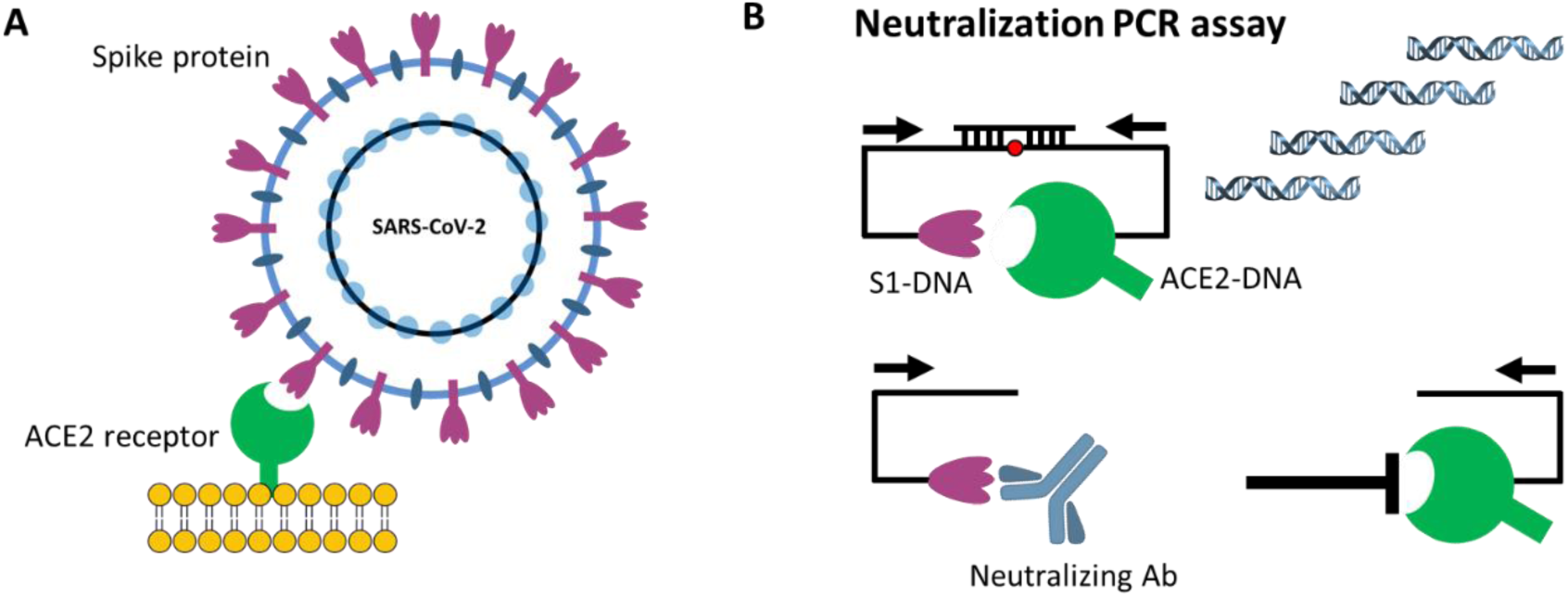
Principle of neutralization PCR assay. (A) Viral entry of SARS-CoV-2 is mediated by the binding of the spike protein to the human receptor angiotensin-converting enzyme 2 (ACE2). Disruption of this interaction forms the basis of neutralizing antibodies (NAbs). (B) Neutralization PCR assay reconstructs this interaction using a pair of S1 subunits of the spike protein- and ACE2-DNA conjugates. In the absence of NAbs, S1 and ACE2 engage with strong affinity, thereby positioning the two DNA barcodes in close proximity for subsequent ligation and PCR-amplification. On the other hand, binding of NAbs blocks S1 from ACE2, leaving the two DNA barcodes separated. Since each barcode has only one PCR primer binding site, they cannot be amplified. Therefore, the quantities of NAbs are correlated with the decrease of PCR amplicon formation.

### Pathogen inactivation process for convalescent plasma samples

The Amotosalen/UV light pathogen reduction system (INTERCEPT, Cerus Corporation, Concord, CA) was used to inactivate potential pathogens in convalescent plasma samples according to the manufacturer’s manual^11,12^. Briefly, the plasma samples were first mixed with amotosalen, then passed through an illumination chamber for UVA light treatment. The amotosalen specifically cross-links nucleic acids within the plasma but leaves the protein component intact. The amotosalen was then removed by flow through a chemical absorption device. The final pathogen-inactivated plasma was stored in a sterile plastic container.

## RESULTS

### Development of a cell-free assay to measure NAbs

Neutralizing antibodies (NAbs) against SARS-CoV-2 are predominantly directed against the receptor binding domain (RBD) of the spike protein where they act to disrupt its interaction with the ACE2 receptor on the human cell surface^13-15^

We thus sought to recreate this competition between the NAb and ACE2 receptors for the spike protein in an in vitro assay. While it is possible to develop such assays using ELISA or other solid-phase assay approaches^7^, the ACE2 receptor is highly conformational and difficult to express. The deposition of ACE2 on solid supports could potentially denature or mask critical epitopes. This would create a departure from their true conformations on viral and cell surfaces. To mitigate this concern, we constructed a solution-phase assay wherein a full-length DNA barcode is split into two halves – one installed on the spike protein, and the other installed on the ACE2 receptor (**Fig. 1**). In the absence of NAbs, the spike protein and the ACE2 receptor naturally engage, positioning the two DNA barcodes in close proximity. Then, the addition of a DNA ligase reunites the two barcodes into a full-length DNA amplicon for amplification and quantification. In contrast, NAbs present in a sample will bind onto the spike protein DNA conjugate and prevent its interaction with the ACE2 receptor conjugate. In this scenario, attenuation of signal is observed, since the two DNA barcodes cannot come into the close proximity for ligation and amplification. Given that there is no need for washing or centrifugation to remove the DNA-barcoded probes, the entire assay is conducted in the solution phase, thus preserving the native antigen conformation.

In the development process, we first evaluated assay performance using the S1 portion of the spike protein versus the receptor binding domain (RBD) fragments of the S1 protein. We assayed two convalescent COVID19 patients and four control specimens from healthy blood donors collected prior to the outbreak (**Fig. 2**). The COVID-19 samples had been analyzed using a cell-based pseudovirus neutralization assay^19^ and confirmed to contain high titers of NAbs. For both antigens, we observed no competition signals from the negative control specimens, and strong competition signals from the COVID-19 samples, indicating effective competition and neutralization of the S1-ACE2 interaction. Observing much stronger signals in the S1 protein-based neutralization-PCR assay (**Fig. 2**), we chose to proceed with the S1 protein for further validation. A possible explanation for this observation is that NAbs may neutralize by binding an adjacent epitope on the S1 protein outside of the RBD but still block binding to ACE2 through steric hindrance as evident in Middle East Respiratory Syndrome (MERS) related NAbs^16^.

**Figure 2.**
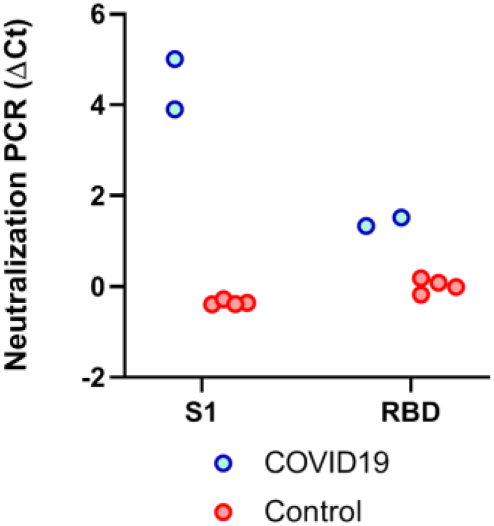
Selection of antigens for neutralization PCR assay. Both full S1 and receptor binding domain (RBD) of S1 have the ability to engage with the ACE2. To evaluate suitability for neutralization PCR assay, we tested convalescent COVID19 patient sera (N=2, blue) and healthy donor sera (N=4, red) collected prior to the outbreak. Notably, the COVID19 sera were confirmed to harbor NAbs by pseudo-virus neutralization assay. The y-axis is the neutralization PCR assay signal ΔCt calculated by subtracting the Ct value of the sample from that of a buffer only blank control.

### Evaluation of neutralization PCR assay sensitivity and specificity

To further validate the assay, we tested blood samples from 18 COVID-19 patients collected 12 days after symptom onset and control specimens collected prior to the outbreak from 43 patients with other infections, including HCV, flavivirus and Lyme disease using the S1-protein neutralization PCR assay (**Fig. 3**). NAbs were detected in all 18 COVID-19 samples but not in any of the 43 non-COVID-19 samples, indicating effective competition and neutralization of the ACE2-S1 interaction. The results demonstrated that this DNA-barcoding assay could detect disruption of S1-ACE2 interaction by NAbs, and, critically, that the NAb signals are specific for SARS-CoV-2.

**Figure 3.**
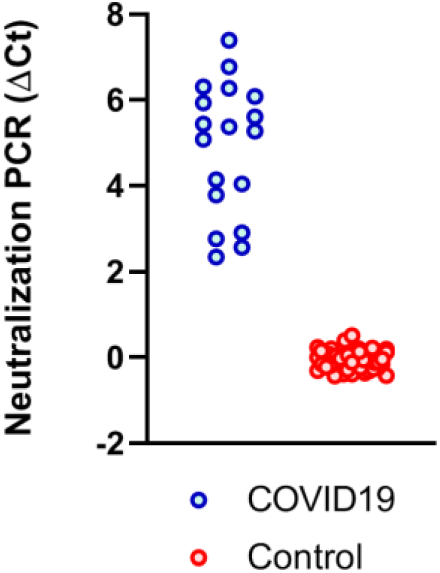
NAbs reactivity in convalescent COVID-19 and control sera with other infection by neutralization PCR assay. Convalescent COVID-19 sera (N=18) were from patients 14 days after symptom onset (blue), and control sera (N=43) from patients with HCV, flavivirus infection and Lyme disease (red).

### Analysis of NAbs development over time

After creating a functional NAb assay, we then analyzed 5 samples from COVID- 19 patients collected within 10 days post symptoms onset, 5 samples between 10- 12 days, 6 samples between 13-15 days, 5 samples between 16-18 days and 7 samples 18 days after symptom onset. The NAb signals quickly increased between 10-12 days and reached a plateau 18 days after onset (**Fig. 4**).

**Figure 4.**
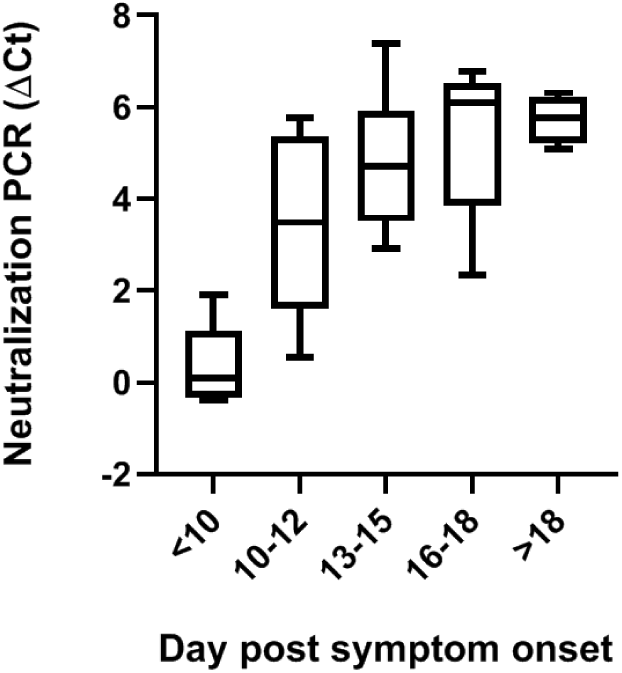
Evaluation of kinetics of NAb development by neutralization PCR assay. Sera from COVID-19 patients after different days post symptom onset were tested by neutralization PCR assay. All sera samples (N=28) were from unique individuals. The boxplots show medians (middle line) and third and first quartiles (boxes), while the whiskers show minimum and maximum above and below the box.

### Evaluation of the impact of CP inactivation protocols on the NAbs

Transfusion of convalescent plasma (CP) for COVID-19 patients may occur in severely ill patients, who are especially vulnerable to the impact of other potentially infectious pathogens in the CP^17^. While donor screening and NAT screening can be implemented to rule out the majority of common blood-borne infectious agents, pathogen reduction treatment for CP is warranted to reduce the risk due to any remaining pathogens not detected by standard blood donor screening tests.

Pathogen inactivation using amotosalen and UVA radiation induced nucleic acid- psoralen adducts has demonstrated efficacy in substantially reduce pathogen burdens in CP^11,12^. We tested samples of CP from three COVID-19 donors before and after pathogen reduction (**Fig. 5**). The NAb signals were highly preserved, indicating retention of the active NAb component after such pathogen reduction treatment.

**Figure 5.**
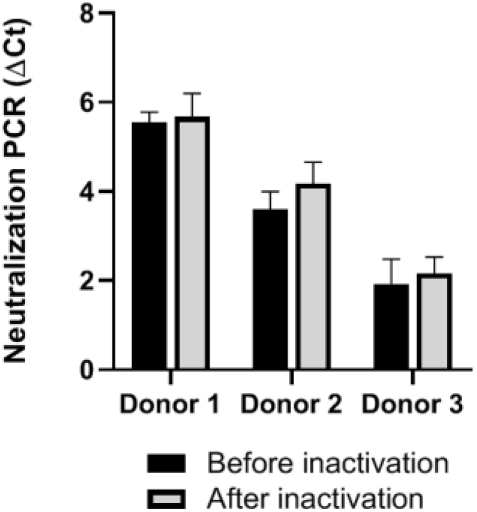
Evaluation of the integrity of NAbs before and after pathogen inactivation treatment of plasma. Three convalescent plasma specimens from recovered COVID-19 patients were treated with amotosalen and UVA light for photo-chemical pathogen inactivation prior to administration to symptomatic COVID-19 patients. The impact on NAbs integrity was evaluated by neutralization PCR assay by comparing signals before (black) and after (gray) pathogen inactivation treatment. Error bars show standard deviations of three replicates.

## DISCUSSION

Analytical techniques for SARS-CoV-2 are essential for a variety of purposes. RNA-based tests detect actively infected patients who might be shedding infectious virus. Serological tests are required to measure the presence of antibodies to aid in understanding individual exposure and prevalence. Neutralizing antibody assays are required to provide insight into immunity of individuals and to recruit CP donors to help treat critically ill patients or to act as short-term immunoprotection for front-line health care workers and immunocompromised and high mortality risk individuals^3^.

We developed an easily deployable test that used standard real-time quantitative PCR (qPCR) to quantify these critical NAbs. The method has the benefit of operational simplicity in that it only requires serial addition of key reagents without washing, centrifugation or cell culturing. The assay can be completed in as little as 140 min and is amenable to high-throughput implementation on laboratory liquid handling workstations. The use of purely recombinant proteins helps standardize assay protocols and results across laboratories. The assay also has the advantage of consuming a very small amount of ACE2 protein per assay, since ACE2 protein is highly costly due to low yield.

Our study revealed that both the RBD and S1 proteins can be used as antigens for our neutralization assay. However, the S1 proteins showed stronger NAb signals. Past MERS studies have identified highly potent neutralizing monoclonal antibodies targeting regions outside of the RBD^16^. The finding warrants future study to investigate whether use of the full-length spike protein (S1+S2) can further enhance NAb detection. It also remains to be evaluated if competition NAb assay signals using any of these antigens correlate with the therapeutic efficacy of the sample.

Furthermore, the study showed that the average NAb signals continued to rise after onset of initial symptoms and then plateau after day 18. These data are consistent with an independent study using the pseudovirus neutralizing assay^14^. This evidence further affirms that cell-free assay formats can obtain results that are concordant with cell-based assays.

Notably, this assay may be used to investigate and qualify different pathogen inactivation procedures for CP. As an example, we showed that UVA/amotosalen treatment did not disrupt the functional integrity of NAbs. Similar retention of antibody neutralization efficacy after pathogen reduction has been reported for convalescent Ebola virus convalescent plasma^18^.

In conclusion, we report development of an easily adoptable assay format to facilitate the measurement of neutralizing function of SARS-CoV-2 antibodies. This assay can complement traditional serological assays by identifying promising CP donors and help focus further analysis on those high-quality donors that merit further characterization using more sophisticated methods including the live virus plaque reduction neutralization test (PRNT). This testing scheme may facilitate a more rapid turn around and delivery of well-characterized convalescent plasma to patients when it is most needed. Future studies are planned to verify assay performance in larger cohorts to broaden the observed correlation with PRNT and to assess utility in terms of improved CP treatment outcomes. Finally, accurate assessment of neutralization capacity is an essential measure of vaccine efficacy. This assay has the potential to assist and accelerate worldwide efforts at developing and validating viable vaccines for SARS-CoV-2.

## Data Availability

The datasets and reagents generated and analyzed during the current study are available from the corresponding author upon reasonable request.

## ACKNOWLEDGEMENTS

We are sincerely grateful to all patients, donors and staff who participated in this study. The study is funded in part by NIH SBIR 2R44DK110005-02 and 2RAI141118- 02 to Enable Biosciences. The content is solely the responsibility of the authors and does not necessarily represent the official views of the NIH.

